# Characterizing the Relationship between Personality Dimensions and Psychosis-Specific Clinical Characteristics

**DOI:** 10.1101/2024.06.06.24308169

**Authors:** Youjin Jenny Jang, Walid Yassin, Raquelle Mesholam-Gately, Elliot S. Gershon, Sarah Keedy, Godfrey G. Pearlson, Carol A. Tamminga, Jennifer McDowell, David A. Parker, Kodiak Sauer, Matcheri S. Keshavan

## Abstract

**Background:** Past studies associating personality with psychosis have been limited by small nonclinical samples and a focus on general symptom burden. This study uses a large clinical sample to examine personality’s relationship with psychosis-specific features and compare personality dimensions across clinically and neurobiologically defined categories of psychoses.

**Methods:** A total of 1352 participants with schizophrenia, schizoaffective disorder, and bipolar with psychosis, as well as 623 healthy controls (HC), drawn from the Bipolar-Schizophrenia Network for Intermediate Phenotypes (BSNIP-2) study, were included. Three biomarker-derived biotypes were used to separately categorize the probands. Mean personality factors (openness, conscientiousness, extraversion, agreeableness, and neuroticism) were compared between HC and proband subgroups using independent sample t-tests. A robust linear regression was utilized to determine personality differences across biotypes and diagnostic subgroups. Associations between personality factors and cognition were determined through Pearson’s correlation. A canonical correlation was run between the personality factors and general functioning, positive symptoms, and negative symptoms to delineate the relationship between personality and clinical outcomes of psychosis.

**Results:** There were significant personality differences between the proband and HC groups across all five personality factors. Overall, the probands had higher neuroticism and lower extraversion, agreeableness, conscientiousness, and openness. Openness showed the greatest difference across the diagnostic subgroups and biotypes, and greatest correlation with cognition. Openness, agreeableness, and extraversion had the strongest associations with symptom severity.

**Conclusions:** Individuals with psychosis have different personality profiles compared to HC. In particular, openness may be relevant in distinguishing psychosis-specific phenotypes and experiences, and associated with biological underpinnings of psychosis, including cognition. Further studies should identify potential causal factors and mediators of this relationship.

## Introduction

The relationship between personality and psychopathology has long been a subject of research and debate, with emphasis on its relevance to general symptom burden and coping (Lysaker, Bell, Kaplan, Greig & Bryson, 1999; Djordjevic et al., 2022), and a burgeoning interest in neurobiological correlates and genetic associations of personality dimensions (Blain et al., 2019; Ohi et al., 2021). Still, there is a limited understanding of personality’s contribution to psychopathology, and the effort to further describe and understand the potential relationship between the two has been ongoing. In most clinical investigations, the five factor model (FFM) of personality—also known as the “Big Five”—has in particular been prioritized for its validity (Costa & McCrae, 1992; Roberts & Delvecchio, 2000), cross-cultural applicability (Allik 2005), and stability independent of state changes (Kentroes et al., 1997, Boyette, Nederlof, Meijer, de Boer & de Haan, 2015).

Efforts to characterize the relationship between personality and psychosis have had a varied course. According to a recent review of literature by Franquillo et al. (2021), the most commonly replicated association between personality and psychosis are high neuroticism, low extraversion, and low agreeableness (Berenbaum & Fujita, 1994; Camisa et al., 2005; Herrán et al., 2006, Reno, 2004). In particular, neuroticism has been implicated heavily in clinical presentations of schizophrenia. Gleeson et al. (2005) found that those who are more likely to relapse after a first episode of psychosis had higher neuroticism and lower agreeableness; Lysaker and Taylor (2007) reported that higher neuroticism and lower extraversion are related to greater emotional discomfort and avoidant coping. Lung, Shu, and Chen (2009) found that patients with neuroticism were more likely to interpret their auditory hallucinations as malevolent, thus increasing distress. These studies have supported the theory that high neuroticism in particular may be a risk factor for the development of schizophrenia and worse outcomes and, conversely, that low neuroticism may be protective (van Os & Jones, 2001; Krabbendam et al. 2002; Boyette et al., 2013).

Other personality traits have shown weaker or more inconsistent relationships with psychotic disorders, though generally higher symptom burden has been associated with lower conscientiousness, extraversion, and agreeableness (Boyette et al., 2014; Franquillo et al., 2021). Openness has been more challenging to characterize. In some investigations, it has been associated with schizotypy and psychoticism in a nonclinical sample (Camisa et al., 2005; DeYoung et al., 2005; DeYoung & Gray, 2009) or subclinical symptoms in probands (Boyette et al., 2013). In another study, openness was shown to be the only variable with no significant relationship to clinical outcomes including duration of untreated psychosis, functioning, or positive and negative symptoms (Compton et al., 2015). Furthermore, another investigation with a small clinical sample, openness to experience was negatively correlated with duration of untreated psychosis, suggesting a possible protective effect (Maric et al., 2018).

Given the enduring nature of personality across one’s lifespan (McCrae, 2009) and their potential heritability (Bouchard & McGue, 2003), each person’s unique constellation of personality dimensions can offer meaningful information about how they respond to their environment and illness burden. Clarifying the potential interactions between personality and psychopathology can therefore have clinical utility: they can offer etiological clues, preventively identify vulnerabilities, or contribute to prognostications (Andersen & Bienvenu, 2011). This can be especially so in chronic and progressive illnesses like primary psychotic disorders, wherein the individual will have to face chronic stressors and would benefit greatly from engagement with treatment and community support.

Past studies that have characterized personality in subjects with psychosis have had two primary limitations: a small or nonclinical sample, and a focus on general symptom burden without specific consideration of psychosis-specific clinical features. This study aims to fill these gaps in the literature.

Using a large and robustly characterized clinical sample collected by the Bipolar and Schizophrenia Network for Intermediate Phenotypes 2 (BSNIP-2) consortium, the study investigates whether personality profiles differ significantly between healthy controls and the psychosis proband. The study explores whether these differences vary across three DSM diagnostic subgroups—schizophrenia (SZ), schizoaffective disorder (SAD), and bipolar disorder with psychosis (BP). Additionally, the study compares personality traits across “Biotypes”, subcategories of psychoses based on biomarkers, distinguishing between biological and clinical categories of psychosis (Clementz et al., 2016).

Further, the study examined personality’s relationship with two principal clinical domains. First is cognition, an important feature of primary psychotic disorders; global cognitive impairment is a key feature of primary psychotic disorders, and a significant contributor to functional outcomes (Mesholam-Gately et al., 2009). Secondly, the association between personality and psychosis-specific symptom burden were explored, including positive symptoms, negative symptoms, and general functioning.

Given the abundance of evidence associating neuroticism with psychosis, we initially hypothesized that neuroticism is the trait most likely to be different between healthy controls and the proband, and most associated with psychotic symptom burden. On the other hand, recent observations suggest a relationship between openness and creativity, mental flexibility and their neural correlates (Abu Raya et al., 2023). For this reason, we hypothesized that cognition specifically may be more related to openness, a personality dimension that includes a facet assessing general intellect.

## Materials and Methods

### Participants

The BSNIP-2 sample was recruited through data collection sites in five cities across the United States (Athens GA, Boston MA, Chicago IL, Dallas TX, and Hartford CT). The study included clinically stable, outpatient participants (n=1352) with diagnoses of SZ (n=560), SAD (n=502), or BPP (n=290) based on the Diagnostic and Statistical Manual of Mental Disorders, Revised 4th Edition (DSM-IV-TR), as well as 623 healthy controls (HC). Exclusion criteria for enrollment included positive tests for controlled substances and head injuries leading to loss of consciousness for longer than 30 minutes (lifetime). Further details on study design and recruitment have been previously described in Tamminga et al. (2013). All participants provided written informed consent approved by each site’s Institutional Review Board. Demographic information for the study population is included in table 1.

**Table 1.** Population Characteristics (Age, Sex, and Race).

|  | Proband | SZ | SAD | BP | BT1 | BT2 | BT3 | HC |
| --- | --- | --- | --- | --- | --- | --- | --- | --- |
| **N** | 1366 | 560 | 500 | 290 | 339 | 342 | 361 | 623 |
| **Age (mean, STD)** | 38.63, 18.941 | 39.69, 25.862 | 39.54, 11.897 | 35.28, 11.487 | 38.50, 12.200 | 40.74, 10.961 | 36.44, 12.237 | 34.37, 12.030 |
| **Sex (% Male)** | 50.1 | 61.1 | 43.4 | 39.7 | 61.1 | 40.6 | 49.6 | 38.4 |
| **Race** |  |  |  |  |  |  |  |  |
| AA | 41.1 | 51.8 | 40.2 | 22.4 | 49.4 | 46.5 | 26.6 | 32.7 |
| CA | 44.8 | 34.8 | 43 | 66.6 | 34.7 | 37.4 | 59.8 | 48.2 |
| OTH | 14.1 | 13.4 | 16.8 | 11 | 15.9 | 16.1 | 13.6 | 19.1 |

Total sample size, mean age (with standard deviation), sex (percent male), and race (% racial group) for each psychosis subgroup and healthy controls, as well as for the aggregate psychosis proband. STD = Standard Deviation; CA=self-identified Caucasian American, AA=self-identified African American, OTH=all other races. SZ=Schizophrenia, SAD=Schizoaffective disorder, BP=bipolar disorder with psychosis, BT1=biotype 1, BT2=biotype 2, BT3=biotype 3, HC=healthy control

## Measures and Rating Scales

### Clinical Assessments

Identical test materials and procedures were used across the five sites and standardized through centralized training. A full clinical characterization of each participant was completed at the time of enrollment. The Structured Clinical Interview for the DSM-IV-TR (SCID) was used to confirm eligible diagnosis, and diagnostic conferences were held across the study sites for consensus.

### Biotype Determinations

The majority of the proband subjects of the BSNIP-2 consortium (1042 of 1350) participated in the characterization of their electrophysical and cognitive biomarkers, including pro- and anti-saccade tasks, auditory paired stimuli and oddball evoked brain responses, the stop signal task, and the Brief Assessment of Cognition in Schizophrenia (BACS) (Clementz et al., 2016). The data was then used to cluster psychosis cases into three distinct biotypes. The patients that formed Biotype-1 (N=339) had poor cognition, with reduced neural responses to salient stimuli; Biotype-2 (N=342) was characterized by impaired cognition and hyper-responsiveness to sensorimotor events; Biotype-3 (N=361) was closest to healthy controls, though still with modest deviations (Tamminga et al., 2017). (See Supplemental Table 1.)

### Personality

Personality was assessed using the self-report questionnaire IPIP-NEO-120 (International Personality Item Pool Representation of the NEO PI-R), based on the five-factor model of personality. The IPIP-NEO-120 contains 120 items which are summed to measure each of the five factors: openness, conscientiousness, extraversion, agreeableness, and neuroticism (Johnson 2014). Each item is scored on a five-point Likert-type scale from 1 to 5. For positively keyed items, the response “strongly disagree” is assigned a value of 1, and “strongly agree,” a value of 5. For negatively keyed items, the scoring is in the opposite direction (“strongly disagree” is assigned a 5, and “strongly agree,” a value of 1). The NEO is the most widely utilized measure of personality with a consistent record of construct validity and internal reliability (Johnson, 2014).

### Cognition

Cognition was assessed using the Brief Assessment of Cognition in Schizophrenia (BACS), an instrument that assesses aspects of cognition most impaired and most strongly correlated with outcomes in patients with schizophrenia (Keefe, 2004). The BACS assesses cognition across six discrete domains: verbal memory, working memory, motor speed, verbal fluency, attention and speed of information processing, and executive functions. The composite score is calculated by averaging the scores from the six measures and calculating a z-score. Only the composite score was used for the analyses in this study.

### Psychotic Symptom Burden

The Positive and Negative Symptom Scale (PANSS) was administered to determine the degree of symptom burden for all participants in the psychosis proband group. PANSS is a comprehensive scale to assess psychopathology and consists of three sub-scales, covering positive symptoms (items P1-P7), negative symptoms (N1-N7), and general symptoms (G1-G16) (Kay, 1987). PANSS items are rated on a 7-point scale from 1 (absent) to 7 (extreme). Based on our *a priori* hypothesis, only the positive and negative subscores were used in this study, to focus on psychosis-specific symptomatology.

### Functional Capacity

The Global Assessment of Functioning (GAF) score is part of the DSM-IV and is the most extensively used method to assess functional impairment for patients with psychosis. The scale ranges from 0 to 100, with 0 representing the most impaired end of the spectrum, and 100 least impaired. Descriptors are provided for each 10-point interval.

### Statistical Analyses

To determine if there are differences in age, sex, and race on personality, Pearson’s correlation coefficients were calculated between age and the five personality factors, and independent sample t-tests were run for sex, and one way Analysis of Variance (ANOVA) for race. Statistical Package for the Social Sciences (SPSS) v28 was used to conduct all statistical analyses.

The means of the five personality factors were compared between healthy controls and the psychosis proband population using independent sample t-tests. This comparison with healthy controls was repeated with each diagnostic and biotype subgroup to see whether the direction of difference was similar across diagnoses. A robust linear regression was used to examine the difference between diagnostic subgroups/biotypes for each personality factor, by incorporating Age, Sex, and Race into the model using the ‘lmrob’ function in R. To ensure robust estimation, ‘MM’ estimator was used as illustrated in Yohai (1987) and Koller and Stahel (2011). Bonferroni correction was used to correct for multiple comparisons. An Analysis of Covariance (ANCOVA) was completed across the diagnostic and biotype proband subgroups to determine main effects of diagnoses and biotypes, and their interactions with age, sex, and race as covariates.

The association between the five personality dimensions and cognition was determined through Pearson’s correlation. Finally, to delineate the relationship between personality traits and clinical features of psychosis, a canonical correlation was run between the five personality factors and psychosis-related variables (positive symptoms, negative symptoms, and functioning). Three primary outcomes of the canonical correlation analysis were reported only in the canonical dimensions that were statistically significant based on a Wilk’s lambda: 1) the correlation coefficient of the two canonical variables, 2) canonical loading coefficients, describing each variable’s contribution to the variance of the canonical variable of its own group, and 3) cross loading coefficients, describing a given variable’s contribution to the variance of the canonical variable of the other group.

## Results

### Participant Data

Significant relationships were found across age in openness (-0.262, p<0.05) and agreeableness (-0.152, p<0.05). There were between-sex differences in conscientiousness (p=0.020), agreeableness (p<0.001), and neuroticism (p<0.001). Race-specific differences were observed in all personality factors (p<0.001 for openness and agreeableness, p<0.05 for conscientiousness and neuroticism), except extraversion. Given these differences, age, sex, and race were included as covariates in subsequent analyses. (See Table 2.)

**Table 2.** Relationship between age, sex and race and the variables of interest.

|  | Age (r, CI) | Sex (t, p) | Race (F, eta sq, p) |
| --- | --- | --- | --- |
| Openness | -0.262 (-0.307, -0.215) | -1.59, 0.119 | 72.19, 0.083, <0.001 |
| Conscientiousness | 0.016 (-0.034, 0.065) | -2.32, 0.020 | 3.298, 0.004, 0.037 |
| Extraversion | -0.152 (-0.199, -0.104) | -0.97, 0.332 | 0.890, 0.001, 0.411 |
| Agreeableness | 0.022 (-0.026, 0.071) | -6.23, <0.001 | 19.447, 0.024, <0.001 |
| Neuroticism | 0.041 (-0.008, 0.090) | -3.89, <0.001 | 4.109, 0.005, 0.017 |
| Cognition | -0.116 (-0.176, -0.055) | 3.08, 0.002 | 69.967, 0.120, <0.001 |

**Mean differences in all variables across age (Pearson correlation), sex (Independent Samples T-Test), race (one-way ANOVA). R=Pearson coefficient for bivariate correlation; CI=95% confidence interval; t=t score for Levene's Test for Equality of; p=level of significance; F=F-statistic for ANOVA; eta sq=measure of effect size.**

### Personality traits

#### Proband v. HC

Significant differences across all five personality traits were observed between the proband and healthy control populations. Overall, the proband population had lower openness, conscientiousness, extraversion, and agreeableness, and higher neuroticism. The magnitude of difference was largest for neuroticism (t=-23.17, p<0.001), followed by conscientiousness (t=18.18, p<0.001), extraversion (t=17.61, p<0.001), agreeableness (t=9.64, p<0.001), and openness (t=2.43, p<0.001). (See Table 3.)

**Table 3.** Comparison of personality factors by subgroup.

|  | t | df | Two-sided p | Mean difference | STE difference | 95% CI (Lower) | 95% CI (Upper) |
| --- | --- | --- | --- | --- | --- | --- | --- |
| **Openness** |  |  |  |  |  |  |  |
| HC-Proband | 2.434 | 1587.000 | 0.015 | 1.589 | 0.653 | 0.308 | 2.869 |
| HC-SZ | 6.483 | 976.000 | <0.001 | 4.655 | 0.718 | 3.246 | 6.064 |
| HC-SAD | 3.203 | 932.000 | 0.001 | 2.569 | 0.802 | 0.995 | 4.143 |
| HC-BP | -6.423 | 765.000 | <0.001 | -6.109 | 0.951 | -7.976 | -4.242 |
| HC-BT1 | 3.415 | 844.000 | <0.001 | 2.870 | 0.840 | 1.220 | 4.520 |
| HC-BT2 | 6.975 | 847.000 | <0.001 | 5.580 | 0.800 | 4.010 | 7.150 |
| HC-BT3 | -3.395 | 883.000 | <0.001 | -2.870 | 0.845 | -4.529 | -1.211 |
| **Conscientiousness** |  |  |  |  |  |  |  |
| HC-Proband | 18.180 | 1584.000 | <0.001 | 13.514 | 0.743 | 12.056 | 14.972 |
| HC-SZ | 13.020 | 978.000 | <0.001 | 11.428 | 0.878 | 9.705 | 13.150 |
| HC-SAD | 17.378 | 934.000 | <0.001 | 15.763 | 0.907 | 13.983 | 17.543 |
| HC-BP | 12.516 | 772.000 | <0.001 | 13.647 | 1.090 | 11.507 | 15.788 |
| HC-BT1 | 14.091 | 848.000 | <0.001 | 13.660 | 0.969 | 11.757 | 15.563 |
| HC-BT2 | 14.489 | 851.000 | <0.001 | 14.220 | 0.981 | 12.294 | 16.146 |
| HC-BT3 | 13.429 | 887.000 | <0.001 | 12.840 | 0.956 | 10.963 | 14.717 |
| **Extraversion** |  |  |  |  |  |  |  |
| HC-Proband | 17.611 | 1587.000 | <0.001 | 14.060 | 0.798 | 12.494 | 15.626 |
| HC-SZ | 15.195 | 979.000 | <0.001 | 13.596 | 0.895 | 11.84 | 15.352 |
| HC-SAD | 17.875 | 933.000 | <0.001 | 16.944 | 0.948 | 15.083 | 18.804 |
| HC-BP | 8.900 | 771.000 | <0.001 | 9.978 | 1.121 | 7.777 | 12.179 |
| HC-BT1 | 12.774 | 849.000 | <0.001 | 13.000 | 1.018 | 11.003 | 14.997 |
| HC-BT2 | 16.783 | 852.000 | <0.001 | 16.370 | 0.975 | 14.456 | 18.391 |
| HC-BT3 | 14.159 | 886.000 | <0.001 | 14.120 | 0.997 | 12.163 | 16.077 |
| **Agreeableness** |  |  |  |  |  |  |  |
| HC-Proband | 9.639 | 1595.000 | <0.001 | 5.694 | 0.591 | 4.535 | 6.852 |
| HC-SZ | 8.728 | 981.000 | <0.001 | 5.905 | 0.677 | 4.578 | 7.233 |
| HC-SAD | 8.557 | 939.000 | <0.001 | 6.127 | 0.716 | 4.722 | 7.532 |
| HC-BP | 5.353 | 771.000 | <0.001 | 4.523 | 0.845 | 2.864 | 6.181 |
| HC-BT1 | 8.215 | 851.000 | <0.001 | 6.2200 | 0.757 | 4.734 | 7.706 |
| HC-BT2 | 9.262 | 853.000 | <0.001 | 7.030 | 0.759 | 5.540 | 8.520 |
| HC-BT3 | 5.446 | 889.000 | <0.001 | 3.960 | 0.727 | 2.533 | 5.387 |
| **Neuroticism** |  |  |  |  |  |  |  |
| HC-Proband | -23.177 | 1588.000 | <0.001 | -19.837 | 0.856 | -21.515 | -18.158 |
| HC-SZ | -15.154 | 976.000 | <0.001 | -15.179 | 1.002 | -17.144 | -13.213 |
| HC-SAD | -23.084 | 937.000 | <0.001 | -23.794 | 1.031 | -25.817 | -21.771 |
| HC-BP | -18.062 | 769.000 | <0.001 | -21.886 | 1.212 | -24.264 | -19.507 |
| HC-BT1 | -17.687 | 846.000 | <0.001 | -19.680 | 1.113 | -21.864 | -17.496 |
| HC-BT2 | -18.881 | 849.000 | <0.001 | -21.010 | 1.113 | -23.194 | -18.826 |
| HC-BT3 | -18.151 | 889.000 | <0.001 | -19.720 | 1.086 | -21.852 | -17.588 |

Independent sample t-tests for mean differences in all five personality factors between each psychosis subgroup and HC, as well as the aggregate psychosis proband and HC. t=t-score from independent sample t-test; df=degree of freedom; Two-sided p=significance of t-test; mean difference=absolute value of difference between means of two groups; SE of difference=standard error of mean difference; 95% CI=95% Confidence Interval for mean difference.

#### Interaction Between Diagnostic Groups v. Biotypes

There was no statistically significant interaction between the biotypes and diagnostic groups in any of the personality traits (p for openness=0.956, conscientiousness=0.153, extraversion=0.869, agreeableness=0.759, neuroticism=0.691).

#### Diagnostic Groups v. HC

The same pattern of personality differences observed between the proband and HC were noted when looking specifically at the SZ and SAD subgroups vs HC. (See Table 3.) In the comparison between BP and HCs, the difference in openness was in the opposite direction; participants with BP tended to have higher openness compared to healthy controls (t=-6.42, p<0.001). (See Table 3.)

#### Biotype Groups v. HC

Personality differences between the biotype 1 subgroup and HC, as well as between the biotype 2 subgroup and HC, were consistent with differences between proband and HC populations (See Table 3.) The biotype 3 subgroup deviated from this trend only in that they had higher openness compared to healthy controls (t=-3.40, p<0.001). (See Table 3.)

#### BP v. SAD v. SZ

When comparing across all subgroups, there was a significant difference in all traits except agreeableness. The largest difference was observed in openness (SZ < SAD < BP, SAD vs. SZ (coefficient estimate (CE) = -1.62, p = 0.136); SAD vs. BP (CE = -6.07, p = 1.16e-07); BP vs. SZ (CE = -7.68, p = 1.46e-12)), followed by neuroticism (SZ < BP < SAD, SAD vs. SZ (CE = - 7.46, p = 2.96e-03); SAD vs. BP (CE = 2.70, p = 0.19); BP v. SZ (CE=-4.77, p=4.05e-09)), extraversion (SAD < SZ < BP, SAD v. SZ (CE=2.31, p=0.14); SAD v. BP (CE=-6.60, p=6.51e-05); BP v. SZ (CE=-4.29, p=0.012)), and conscientiousness (SAD < BP < SZ, SAD vs. SZ (CE = 4.58, p = 4.68e-05); SAD vs. BP (CE = -3.37, p = 0.030); BP vs. SZ (CE = 1.22, p = 1.07)).

Additionally, age was associated with openness (p=2e-16), extraversion (p=5.48e-4), and agreeableness (p=0.0046). Sex was significantly associated with extraversion (p=0.0364), agreeableness (p=9.13e-03), and neuroticism (p=2.37e-09). Race was significantly associated with all traits except extraversion. (See Table 4 for detailed results.)

**Table 4.** Personality across diagnostic and biotype subgroups (with age, sex, race as covariates) Diagnostic Subgroups (DSM Categories)

|  |  | Coefficient<br>t | St. Error | p | R sq. (%) |
| --- | --- | --- | --- | --- | --- |
| Openness | BP v. SAD | -6.066 | 1.095 | 1.16E-07 | 25.08 |
|  | BP v. SZ | -7.684 | 1.049 | 1.46E-12 |  |
|  | SAD v. SZ | -1.617 | 0.807 | 0.1362 |  |
| Conscientiousness | BP v. SAD | -3.369 | 1.305 | 0.02997 | 2.488 |
|  | BP v. SZ | 1.215 | 1.322 | 1.074 |  |
|  | SAD v. SZ | 4.584 | 1.056 | 4.68E-05 |  |
| Extraversion | BP v. SAD | -6.599 | 1.547 | 6.51E-05 | 3.589 |
|  | BP v. SZ | -4.289 | 1.481 | 0.01155 |  |
|  | SAD v. SZ | 2.309 | 1.152 | 0.1359 |  |
| Agreeableness | BP v. SAD | -1.134 | 1.052 | 0.846 | 4.523 |
|  | BP v. SZ | 0.154 | 1.067 | 2.655 |  |
|  | SAD v. SZ | 1.288 | 0.875 | 0.423 |  |
| Neuroticism | BP v. SAD | 2.696 | 1.453 | 0.1914 | 8.752 |
|  | BP v. SZ | -4.766 | 1.443 | 2.96E-03 |  |
|  | SAD v. SZ | -7.462 | 1.220 | 4.05E-09 |  |

**Biotype Subgroups**
|  |  | Coefficient<br>t | St. Error | p | R sq. (%) |
| --- | --- | --- | --- | --- | --- |
| Openness | BT1 v. BT2 | -2.122 | 0.899 | 0.0555 | 22.63 |
|  | BT1 v. BT3 | 3.484 | 1.030 | 2.26E-03 |  |
|  | BT2 v. BT3 | 5.606 | 0.970 | 3.06E-08 |  |
| Conscientiousness | BT1 v. BT2 | -0.820 | 1.224 | 1.509 | 0.772 |
|  | BT1 v. BT3 | 1.381 | 1.234 | 0.789 |  |
|  | BT2 v. BT3 | 2.201 | 1.244 | 0.2313 |  |
| Extraversion | BT1 v. BT2 | -2.999 | 1.387 | 0.0927 | 1.781 |
|  | BT1 v. BT3 | -1.528 | 1.429 | 0.855 |  |
|  | BT2 v. BT3 | 1.471 | 1.324 | 0.801 |  |
| Agreeableness | BT1 v. BT2 | -1.695 | 0.982 | 0.2547 | 6.207 |
|  | BT1 v. BT3 | 1.115 | 0.925 | 0.684 |  |
|  | BT2 v. BT3 | 2.809 | 0.955 | 0.01008 |  |
| Neuroticism | BT1 v. BT2 | 0.340 | 1.423 | 2.433 | 4.392 |
|  | BT1 v. BT3 | -1.298 | 1.412 | 1.074 |  |
|  | BT2 v. BT3 | -1.638 | 1.415 | 0.741 |  |

**Covariates: Personality and Age, Sex, Race**
|  |  | Coefficient | St. Error | p |
| --- | --- | --- | --- | --- |
| Openness | Age | -0.270 | 0.0308 | 2.00E-16 |
|  | Sex (M) | 0.447 | 0.727 | 0.539 |
|  | Race (CA) | 7.071 | 0.8546 | 2.00E-16 |
|  | Race (OTH) | 3.990 | 1.098 | 2.94E-04 |
| Conscientiousness | Age | 0.0746 | 0.0410 | 0.0695 |
|  | Sex (M) | -0.0116 | 0.946 | 0.99 |
|  | Race (CA) | -2.263 | 1.053 | 0.032 |
|  | Race (OTH) | 0.898 | 1.428 | 0.53 |
| Extraversion | Age | -0.160 | 0.0462 | 5.48E-04 |
|  | Sex (M) | 2.198 | 1.049 | 0.0364 |
|  | Race (CA) | -1.554 | 1.146 | 0.175 |
|  | Race (OTH) | 0.260 | 1.624 | 0.873 |
| Agreeableness | Age | 0.0923 | 0.0325 | 4.62E-03 |
|  | Sex (M) | -2.046 | 0.783 | 9.13E-03 |
|  | Race (CA) | 5.273 | 0.859 | 1.20E-09 |
|  | Race (OTH) | 3.17 | 1.199 | 8.37E-03 |
| Neuroticism | Age | -0.0134 | 0.0460 | 0.771 |
|  | Sex (M) | -6.413 | 1.065 | 2.37E-09 |
|  | Race (CA) | 2.584 | 1.161 | 0.0262 |
|  | Race (OTH) | 1.578 | 1.716 | 0.358 |

Coefficient = estimated coefficient; St. error = standard error; p = p value.

#### BT1 v. BT2 v. BT3

Across the three biotypes, there was a significant difference across openness (BT2 < BT1 < BT3, BT1 v. BT2 (CE = 2.12, p = 0.0555); BT1 v. BT3 (CE =3.48, p = 2.26e-03); BT2 v. BT3 (CE = 5.61, p = 3.06e-08)) and agreeableness (BT2 < BT1 < BT3, BT1 v. BT2 (CE = -1.695, p = 0.2547); BT1 v. BT3 (CE = 1.115, p =0.684); BT2 v. BT3 (CE = 2.809, p = 0.01008)). (See Table 4 for detailed results.)

### Personality dimensions and cognition

Of the five personality traits, openness (R=0.383, p<0.001) and agreeableness (R=0.102, p<0.001) were significantly positively correlated with cognition. (See Table 5.)

**Table 5.** Pearson’s Correlation between Personality Dimensions and Cognition.

|  | R | Sig |
| --- | --- | --- |
| Openness | 0.383 | <0.001 |
| Conscientiousness | 0.037 | 0.265 |
| Extraversion | 0.033 | 0.324 |
| Agreeableness | 0.102 | 0.002 |
| Neuroticism | -0.002 | 0.946 |

#### Canonical correlation

Two of the three canonical correlations between the five personality traits and psychosis-specific symptoms were significant, with a correlation value of 0.36 (p<0.001) for the first dimension, and 0.17 (p<0.001) for the second dimension (Fig. 1).

**Figure 1.**
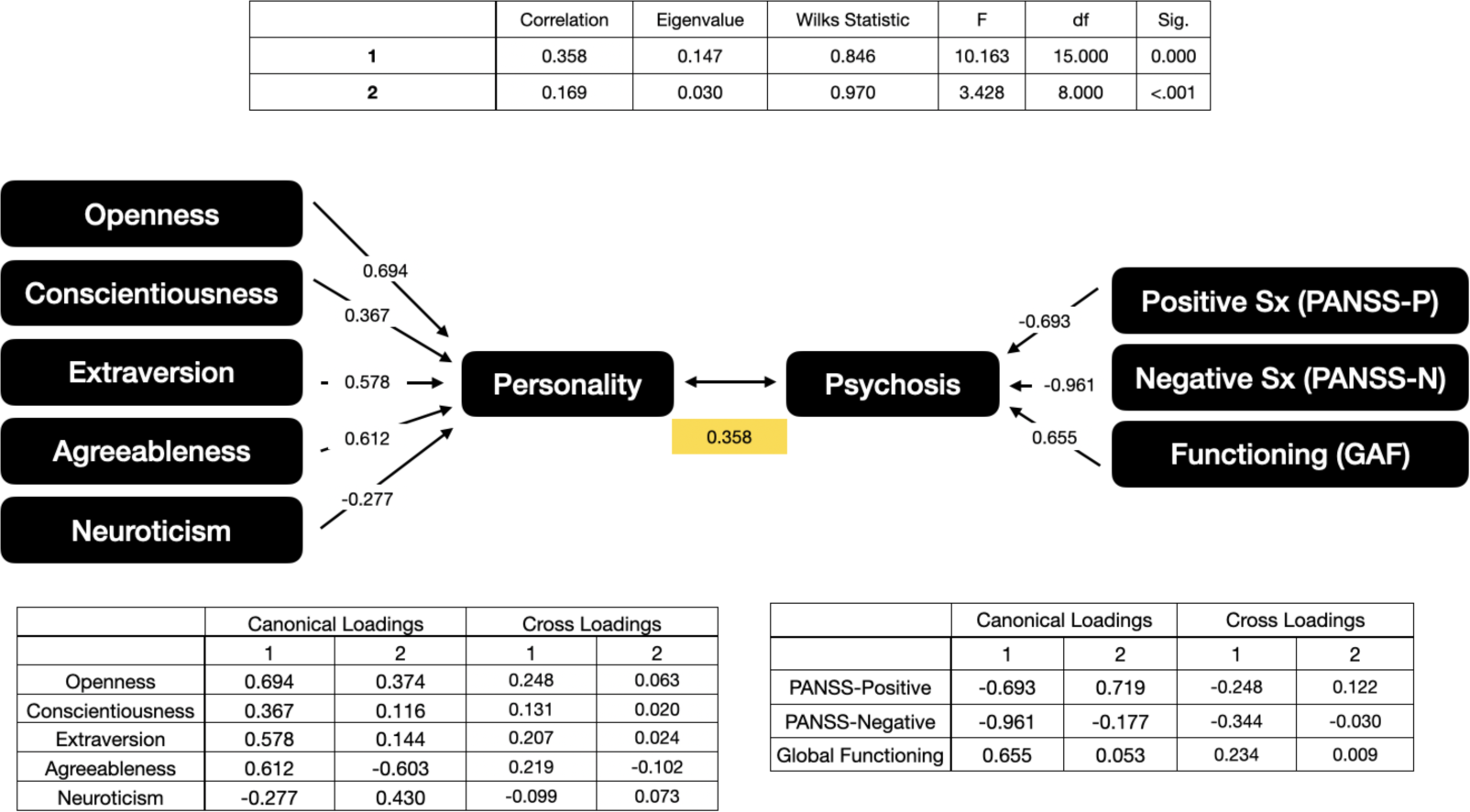
Canonical Correlation (personality vs. psychosis variables) Canonical correlation between the five personality factors and psychosis-related outcome variables (positive symptoms, negative symptoms, global functioning) for the psychosis proband. Correlation=Pearson correlation coefficient of the two canonical functions; F=F-statistic associated with the degree of freedom; df= Degree of freedom; Sig.=significance of canonical correlation by Wilk’s Lambda test.

For the first canonical dimension, the personality factors with the highest cross-loading values were openness (0.25), agreeableness (0.22), and extraversion (0.21), followed by conscientiousness (0.13) and neuroticism (-0.10). The psychosis-specific variables with the highest cross-loading were negative symptoms (-0.34), positive symptoms (-0.25), and functioning (0.23).

For the second canonical dimension, the personality factors with the highest cross-loading were agreeableness (-0.10), neuroticism (0.07), and openness (0.06), followed by extraversion (0.02) and conscientiousness (0.02). The psychosis-specific variables with the highest cross-loading were positive symptoms (0.12), negative symptoms (-0.03), and functioning (0.01).

## Discussion

This study was aimed at delineating the relationship between personality and psychosis by describing whether patients with psychotic disorders have different personality traits compared to healthy controls and illustrate the relationship between these personality traits and psychosis-related symptom burden. Our findings showed that there were significant personality differences between the psychosis proband and healthy controls across all five personality domains; overall, healthy controls had lower neuroticism and higher extraversion, agreeableness, conscientiousness and openness. The lack of interaction effect suggests that the effect of biotype and diagnosis on personality may be independent of each other, i.e., they are not interacting with each other to significantly influence the personality factors. Thus the effect of biology on personality may not depend on the diagnosis, and vice versa. Openness was the trait with the most significant difference across the diagnostic subgroups and biotypes: compared to HC, BP and BT3 had higher openness, while SAD, SZ, BT1, and BT2 subgroups had lower openness. Regarding personality’s relationship with cognition, openness was shown to be the personality trait with the strongest association. As for symptom burden, a canonical correlation analysis between the five personality traits and psychosis-specific symptom burden showed that openness, agreeableness, and extraversion had the strongest association with symptom severity.

The results from personality comparisons between the control and the proband populations aligns with and supports existing literature that individuals with psychosis tend to have higher neuroticism and lower conscientiousness, extraversion, agreeableness, and openness compared to healthy controls (Boyette et al., 2015). As our results and those of others suggest, it is conceivable that higher neuroticism predisposes an individual to greater anxiety, aggravating the distress or interpretation of psychotic thought content (van Os et al., 2001, Krabbendam et al., 2002, Boyette et al., 2013). It could also be speculated that lower conscientiousness, extraversion, and agreeableness might contribute to the alienating impact of psychosis by making it more challenging to sustain an extensive social network, employment, or routines of self-care (Franquillo et al., 2021).

A key novel finding of this study is the significance of the openness trait. The fact that openness is the personality dimension with the largest difference across diagnostic subgroups suggests its relevance in distinguishing the illness experiences across the diagnoses. Given that openness is also the only dimension with a significant difference across the three biotypes, it is reasonable to posit that openness may be associated with biological underpinnings of psychosis. Since biotype 3 is the only biotype characterized by near-normal cognition and is the subgroup with the highest openness, the biological connection between the openness trait and psychosis may be in the cognitive domain.

The results of the canonical correlation further draw our attention to the potential significance of openness. The design of our analysis, which focuses on psychosis-specific symptoms rather than general symptom burden, shows that the personality trait that demonstrates the strongest association is openness. This suggests that the trait may be relevant in the development or manifestation of positive and negative symptomatology, and more broadly impact functioning.

The key to understanding these results may lie in a more detailed understanding of the openness trait. The effort to understand openness’ relationship with psychoticism is not new. While the trait has not been strongly correlated in studies focusing solely on symptomatology, its relationship to neurobiological correlates of psychosis have been more noteworthy (Grazioplene et al., 2014; Grazioplene et al., 2016). Clarifying this relationship has been complicated by the fact that openness harbors two somewhat conflicting facets in relation to psychosis: “openness to experience,” which may predispose someone to psychoticism, and “intellect,” which may be protective for clinically significant functional decline (DeYoung, 2015, Olyenick et al., 2017).

Some researchers have tried to divide the trait by these two general principles and distinguish whether the different facets contribute differently to psychosis. Blain and DeYoung found, for example, that openness to experience co-varied with measures of psychoticism and was positively correlated to default mode network, which is implicated in psychosis pathophysiology (Whitfield-Gabrieli et al., 2009). Conversely, the intellect facets were positively correlated with frontoparietal network coherence, which is considered more protective.

While these past studies have been limited by a largely nonclinical population and the limited validity of facet-level personality analyses, understanding openness as a multi-dimensional trait can provide a useful framework for the results from this study. Higher openness to experience, for instance, may be related to positive symptoms, which are prominent (compared to negative symptoms) in bipolar disorder. There have been a few studies that have observed, in particular, a brief increase in the openness trait with hallucinogenic experiences in the context of psychotropic drug use (Maclean et al., 2011; Carhart-Harris et al., 2016). Lower intellect might be more strongly related to negative symptoms, which are more pronounced in SZ or SAD, and shown to be associated with cognitive decline and neocortical reductions (Tamminga et al., 2014, Sheffield et al., 2018). Openness may be thus associated with psychosis in a two-pronged manner, embodying both increased psychoticism and decreased cognition.

This is not to discount the importance of the other traits, including agreeableness and extraversion, or to undermine the significance of neuroticism in the overall disease burden. Indeed, results from this study, wherein neuroticism is the dimension most different between the proband and psychosis, corroborate the trait’s likely heavy contribution to patient distress. However, in focusing our attention to symptoms that characterize psychosis, we may be able to formulate pathways specific to the development and persistence of psychosis.

### Strengths and limitations

With the BSNIP2 dataset, we explored personality differences across the psychosis proband and healthy controls in a large, well-characterized and deeply phenotyped sample. Given that the largest past sample in the literature involved 217 subjects in the proband (Boyette et al., 2013), this study’s proband sample (n=1366) offers significantly greater power in comparison. The well-validated biotype distinctions, based on robust electrophysiological and cognitive data and externally validated by imaging and genetic sampling, further add to the strength of the analyses, though of course the biotypes are as yet validated only within the consortium. Further, the study used a longer and more extensive version of the personality factor assessment (IPIP-NEO-120) compared to the typically utilized 60-item assessment, and thus has the potential to capture the five traits with more granularity.

The study is limited by the cross-sectional nature of the data and cannot make claims of temporality or causality. Further, although personality traits are considered stable across disease states including psychotic experiences, the extent to which state experiences influence reporting of trait characteristics is still largely unknown (Boyette et al., 2015). It is possible that psychosis itself may have an impact on the different personality traits, amplifying existing personality traits. Given these limitations, follow-up studies with longitudinal data would be necessary to substantiate the findings. However, the relationships identified here serve as an important inference for continuing studies. Additionally, given that the study population consists primarily of subjects who are in remission or optimally treated in the outpatient setting, the results are likely to underestimate, rather than overestimate, effect size.

### Future directions

The aim of investigating the associations between personality and psychopathology is to better characterize how aspects of an individual’s stable character can influence elements of illness experience and heighten or alleviate distress, and not to pathologize any particular constellation of personality traits.

In conceptualizing the results, it is also important to remember that personality does not exist in a vacuum. Instead, personality is only one element in a complex behavioral pathway that involves many other environmental, genetic and other factors, including trauma, substance abuse or duration of untreated psychosis (Pos et al., 2016). This study identified personality dimensions that are associated with the experience of psychosis broadly; the next step would be to identify which other factors exist in the pathway, and how these elements interact. Clarifying the influence of other factors can also help identify how to shape treatment. Investigating personality dimensions in clinical and familial high risk studies are also needed to longitudinally investigate whether personality alterations may exist prior to psychosis onset.

## Data Availability

The data is available from the NIMH NDA.

https://nda.nih.gov/edit_collection.html?id=2165

## Acknowledgements

We thank all the participants and staff who are part of the BSNIP 2 consortium. We also thank the National Institute of Mental Health for the funding. Finally, we are grateful to Brett Clementz for his valuable contribution to the study.

---

Supplementary Material (adapted from Tamminga et al., 2017)

Biotype Cluster Characteristics
| Biotype 1 | Biotype 2 | Biotype 3 |
| --- | --- | --- |
| Very low cognition | Low cognition | Nearly normal cognition |
| Very low EEG power | High EEG power | Just low on EEG power |
| Low gray matter volume | N100, P300 (normal) | Fast visual orienting |
| Worst negative symptoms | Low cortical volume, more focused |  |
| Highest affected relatives | High affected relatives | Low % affected relatives |
| High polygene score |  | Normal polygene score |
| Lowest cannabis use | Moderate cannabis use | High cannabis use |

EEG, electroencephalography; NL, normal

